# Immune response and endocytosis pathways are associated with the resilience against Alzheimer’s Disease

**DOI:** 10.1101/19009464

**Authors:** Niccolò Tesi, Sven J. van der Lee, Marc Hulsman, Iris E. Jansen, Najada Stringa, Natasja M. van Schoor, Philip Scheltens, Wiesje M. van der Flier, Martijn Huisman, Marcel J. T. Reinders, Henne Holstege

**Affiliations:** Alzheimer Center Amsterdam, Department of Neurology, Amsterdam Neuroscience, Vrije Universiteit Amsterdam, Amsterdam UMC, Amsterdam, The Netherlands; Department of Clinical Genetics, Amsterdam UMC, Amsterdam, The Netherlands; Delft Bioinformatics Lab, Delft University of Technology, Delft, The Netherlands; Department of Complex Trait Genetics, Center for Neurogenomics and Cognitive Research, Amsterdam Neuroscience, VU University, Amsterdam, the Netherlands; Department of Epidemiology and Biostatistics, Amsterdam UMC, Amsterdam, The Netherlands; Amsterdam Public Health Research Institute, Amsterdam, The Netherlands

**Keywords:** centenarians, immune response, endosomal system, Alzheimer’s disease, pathways, polygenic risk score, PRS, longevity, resilience, cognitive health, etiology

## Abstract

Developing Alzheimer’s disease (AD) is influenced by multiple genetic variants that are involved in five major AD-pathways. Per individual, these pathways may differentially contribute to the modification of the AD-risk. The pathways involved in the *resilience* against AD have thus far been poorly addressed. Here, we investigated to what extent each molecular mechanism associates with (i) the increased risk of AD and (ii) the *resilience* against AD until extreme old age, by comparing pathway-specific polygenic risk scores (pathway-PRS). We used 29 genetic variants associated with AD to develop pathway-PRS for five major pathways involved in AD. We developed an integrative framework that allows multiple genes to associate with a variant, and multiple pathways to associate with a gene. We studied pathway-PRS in the Amsterdam Dementia Cohort of well-phenotyped AD patients (N=1,895), Dutch population controls from the Longitudinal Aging Study Amsterdam (N=1,654) and our unique 100-plus Study cohort of cognitively healthy centenarians who avoided AD (N=293). Last, we estimated the contribution of each pathway to the genetic risk of AD in the general population. All pathway-PRS significantly associated with increased AD-risk and (in the opposite direction) with resilience against AD (except for *angiogenesis, p*<0.05). The pathway that contributed most to the overall modulation of AD-risk was β-amyloid metabolism (29.6%), which was driven mainly by *APOE*-variants. After excluding *APOE* variants, all pathway-PRS associated with increased AD-risk (except for *angiogenesis, p*<0.05), while specifically *immune response* (*p*=0.003) and *endocytosis* (*p*=0.0003) associated with resilience against AD. Indeed, the variants in these latter two pathways became the main contributors to the overall modulation of genetic risk of AD (45.5% and 19.2%, respectively). The genetic variants associated with the resilience against AD indicate which pathways are involved with maintained cognitive functioning until extreme ages. Our work suggests that a favorable immune response and a maintained endocytosis pathway might be involved in general neuro-protection, which highlight the need to investigate these pathways, next to β-amyloid metabolism.

## Introduction

Due to changes in lifestyle and advances in healthcare, life expectancy has greatly increased during the last century.[1] A consequence of an increased fraction of aged individuals in the population is the increased prevalence of age-related diseases. A major contribution to poor health and disability at old age is cognitive decline due to Alzheimer’s disease (AD).[2] The incidence of AD increases exponentially with age and reaches ∼40% per year at 100 years, making it one of the most prevalent diseases in the elderly.[3] Yet, a small proportion of the population (<0.1%) avoids the disease, reaching at least 100 years while maintaining a high level of cognitive health.[4]

Both the development and the resilience against AD are determined by a combination of beneficial and harmful environmental and genetic factors that is unique for each individual.[1, 5, 6] Thus far, large collaborative genome-wide association studies (GWAS) have discovered common genetic variants associated with a small modification of the risk of AD.[7–20] Of these, the alleles that encompass the *APOE* gene explain the largest proportion of the risk to develop or the chance to escape AD. We previously showed that those who avoided cognitive decline until extreme ages (cognitively healthy centenarians) were relatively depleted with genetic variants associated with an increased risk of AD.[21] However, the degree of depletion of these variants in the genomes of cognitively healthy centenarians relative to the middle-aged healthy individuals was not constant, which might point towards a differential impact of associated biological pathways on either avoiding or developing AD. This led us to hypothesize that an individuals’ chance to develop AD or to being resilient against AD may be determined by pathway-specific risk.

Previous studies indicated that five specific biological pathways associate strongly with AD risk: *immune response, β-amyloid metabolism, cholesterol/lipid dysfunction, endocytosis* and *angiogenesis*.[22–27] However, the extent to which different pathways contribute to the polygenic risk of AD is unknown. The degree to which a pathway contributes to the individual risk can be studied with pathway-specific polygenic risk scores (PRS).[28, 29] In a typical polygenic risk score, the effect-sizes of all genetic variants that significantly associate with a trait are combined.[30] In a pathway-specific PRS, additional information is necessary: (*i*) the association of genetic variants to genes, and (*ii*) the association of genes to pathways. Previous studies of pathway-PRS in AD approached these challenges using the closest gene for variant mapping. For this, a 1:1 relationship between variants and genes is assumed, however, as AD-associated variants are mostly intronic or intergenic, the closest gene is not necessarily the gene affected by the variant. Additionally, different databases often have different functional annotations of genes, and this uncertainty was previously not taken into account when constructing pathway-PRS.[28, 29]

An accurate mapping of the genetic risk of AD conferred by specific molecular pathways may lead to a greater comprehension of individual AD subtypes and might represent a first important step for the development of targeted intervention strategies and personalized medicine.[31] Here, we propose a novel integrative framework to construct pathway-PRS for the five major pathways suggested to be involved in AD. We then tested whether specific pathways differentially contributed to the risk of AD as well as to the chance of avoiding AD until extreme old ages. Finally, we estimated the contribution of each pathway to the polygenic risk of AD in the general (healthy middle-aged) population.

## Methods

### Populations

Population subjects are denoted by *P*: they consist of a representative Dutch sample of 1,779 individuals aged 55-85 years from the Longitudinal Aging Study Amsterdam (LASA).[32, 33] Patients diagnosed with AD are denoted by *A*. The patients are either clinically diagnosed probable AD patients from the Amsterdam Dementia Cohort (N=1,630) or pathologically confirmed AD patients from the Netherlands Brain Bank (N=436).[34–36] Escapers of AD are denoted by *C*: these are 302 cognitively healthy centenarians from the 100-plus Study cohort. This study includes individuals who can provide official evidence for being aged 100 years or older and self-report to be cognitively healthy, which is confirmed by a proxy.[4] All participants and/or their legal representatives provided written informed consent for participation in clinical and genetic studies. The Medical Ethics Committee of the Amsterdam UMC (METC) approved all studies.

### Genotyping and imputation

We selected 29 common genetic variants (minor allele frequency >1%) for which a genome-wide significant association with clinically identified AD cases was found (*Table S1*).[7–18, 37–40] We genotyped all individuals using Illumina Global Screening Array (GSAsharedCUSTOM_20018389_A2) and applied established quality control measures.[41] Briefly, we used high-quality genotyping in all individuals (individual call rate >98%, variant call rate >98%) and Hardy–Weinberg equilibrium-departure was considered significant at *p*<1×10^−6^. Genotypes were prepared for imputation using provided scripts (HRC-1000G-check-bim.pl).[42] This script compares variant ID, strand and allele frequencies to the haplotype reference panel (HRC v1.1, April 2016).[35] Finally, all autosomal variants were submitted to the Michigan imputation server (https://imputationserver.sph.umich.edu). The server uses SHAPEIT2 (v2.r790) to phase data and imputation to the reference panel (v1.1) was performed with Minimac3. Variant-genotypes of total of 1,779 population subjects, 302 centenarians and 2,052 AD cases passed quality control. Prior to analysis, we excluded individuals of non-European ancestry (*NC* = 2, *N*_*P*_ = 63 and *N*_*A*_ = 94 based on 1000Genomes clustering)[43] and individuals with a family relation (*N*_*C*_ = 7, *N*_*P*_ = 62 and *N*_*A*_ = 63, identity-by-descent >0.3), leaving 1,654 population subjects, 293 cognitively healthy centenarians and 1,895 AD cases for the analyses.

### Polygenic risk score

To calculate the personal polygenic risk scores, or the genetic risk of AD that affects a single individual, the effect-sizes of all genetic variants that significantly associate with AD are combined. Formally, a PRS is defined as the sum of trait-associated alleles carried by an individual across a defined set of genetic loci, weighted by effect-sizes estimated from a GWAS.[30] We constructed a polygenic risk score (PRS) using 29 variants that were previously associated with AD. As weights for the PRS, we used the variant effect-sizes (log of odds ratio) as published in large GWAS of AD (*Table S1*). Given a subject *s*, the PRS is defined as:

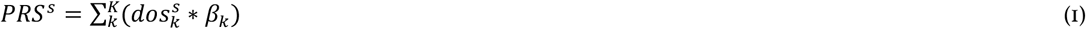

where *K* is the full set of variants, 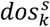 is the allele dosage from the (imputed) genotype of variant *k* in subject *s* and *β*_*k*_ is the effect size as determined in the largest published AD case-control GWAS (*Table S1*).

### Mapping variants to pathways

We studied the five pathways implicated in AD: *immune response, β-amyloid metabolism, cholesterol/lipid dysfunction, endocytosis* and *angiogenesis*.[22–25, 44, 45] For these pathways we developed the *variant-pathway mapping* 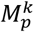, which represents the degree of involvement of a given variant in the pre-selected pathways. To generate this value, we (*i*) associated genetic variants to genes (*variant-gene mapping*), (*ii*) associated genes to pathways (*gene-pathway mapping*) and (*iii*) combined these mappings in the *variant-pathway mapping*.

#### Variant-gene mapping

the association of a variant with a specific gene is not straight-forward as the closest gene is not necessarily the gene affected by the variant. The two most recent and largest GWAS of AD addressed the relationship between genetic variants and associated genes applying two independent methods.[19, 20] Briefly, one study used (*i*) gene-based annotation, (*ii*) expression-quantitative trait loci (eQTL) analyses, (*iii*) gene cluster/pathway analyses, and (*iv*) differential gene expression analysis between AD cases and healthy controls.[19] The other study integrated (*i*) positional mapping, (*ii*) eQTL gene-mapping, and (*iii*) chromatin interaction as implemented in the tool Functional Mapping and Annotation of Genome-Wide Association Studies (FUMA).[20, 46] The list of genes most likely affected by each variant was obtained from both studies and used to derive a weighted mapping for each genetic variant *k* to one or more genes *g*, 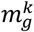, denoted as the *variant-gene mapping* weight. This weight was calculated by counting the number of times a variant *k* was associated with gene *g* across the two studies and dividing this by the total number of genes associated with the variant (*Table S2*). For variants in/near *CR1, PILRA, PLCG2, ABCA7* and *APOE*, we assumed the culprit gene as known, and we assigned a 1:1 relationship between the variant and the gene (*Table S2*).

#### Gene-pathway mapping

each gene from the *variant-gene mapping* was classified into the pre-defined set of pathways integrating four sources of information:

1. Gene-sets from the unsupervised pathway enrichment analysis within MAGMA statistical framework from *Kunkle et al*.,[19] in which the authors identified 9 significant pathways (coupled with the genes involved in each pathway), which we mapped to 3 of the 5 pathways of interest (*Table S3*);
2. Associated genes from Gene-ontology (GO, from *AmiGO 2* version 2.5.12, released on 2018-04) terms resembling the 5 pathways of interest within the biological processes tree (including all child-terms) (*Table S4*);[47, 48]
3. Gene-sets derived from an unsupervised functional clustering analysis within DAVID (v6.8, released on 2016-10):[49, 50] the gene-set from the *variant-gene mapping* was used to obtain 12 functional clusters which were then mapped to the 5 pre-selected pathways using a set of keywords (*Table S5* and *Table S6*);
4. Gene-pathway associations from a recent review concerning the genetic landscape of AD (*Table S7*);[22] By counting the number of times each gene was associated to each pathway according to these sources, and dividing by the total number of associations per gene, we obtained a weighted mapping of each gene *g* to one or more pathways *p*, 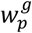, denoted as the *gene-pathway mapping* weight (*Table S8* and *Table S9*). In case the *gene-pathway mapping* could not be calculated (*i*.*e* there was no mapping to any of the pathways of consideration), we excluded the gene from further analyses (*Table S8* and *Table S9*).

To associate variants with pathways, we combined the *variant-gene mapping* and the *gene-pathway mapping*. Given a variant *k*, mapping to a set of genes *G*, and a pathway *p*, we define the weight of the variant to the pathway 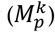 as:

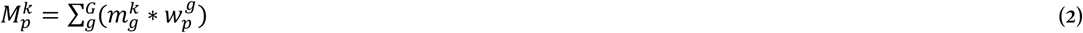

where 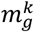 is the *variant-gene mapping* weight of variant *k* to gene *g*, and 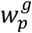 is the *gene-pathway mapping* weight of gene *g* to pathway *p*. In this way, for each variant, we calculated a score indicative of the involvement of the variant in each of the five pathways (*variant-pathway mapping, Table S10*). For some variants no *variant-pathway mapping* was possible. We marked these variants as unmapped (*Table S10*).

### Pathway-specific polygenic risk score

For the pathway-specific polygenic risk score (pPRS), we extended the definition of the PRS by adding as multiplicative factor the *variant-pathway mapping* weight of each variant. Given a sample *s* and a pathway *p*, we defined the pPRS as:

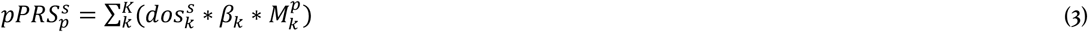

where 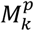 is the *variant-pathway mapping* of variant *k* to pathway *p*.

### Association of PRSs in the three cohorts

We calculated the polygenic risk score (PRS) and pathway-PRS (pPRS) for the population subjects, the AD cases and the cognitively healthy centenarians (*P*, A and *C*, respectively). Prior to analyses, the PRSs of all three populations were combined together and were scaled (mean=0, SD=1). We then investigated the influence of *APOE*, gender and age on the risk scores: we calculated the PRSs and pPRSs with and without the two *APOE* variants and we correlated the resulting (p)PRSs with sex, age (age at inclusion for controls, age at onset for cases) and population substructure components. To inspect the differential contributions of the risk scores to AD development or resilience against AD, we calculated (*i*) the association of the risk scores (PRS and pPRS) with AD status by comparing AD cases and population subjects (*A vs. P*), and (*ii*) the association of the risk scores with resilience against AD by comparing cognitively healthy centenarians and population subjects (*C vs. P* comparison). For the associations, we used logistic regression models with the PRS and pPRS as predictors, adjusting for population substructure (principal components 1-5). Resulting effect-sizes (log of odds ratio) can be interpreted as the odds ratio difference per one standard deviation (SD) increase in the PRS, with a corresponding estimated 95% confidence intervals (95% CI). Association analyses of the (p)PRS in the three population were also stratified by sex. Last, we verified the classification performances of the single variants as well as the (p)PRS by calculating the area under the ROC curve for classification of AD and resilience against AD.

### Comparison of effect-size between *resilience* against AD and increased AD-risk

To further investigate the relationship between the effect of each pathway on AD and on resilience against AD, we calculated the *change* in effect-size. This corresponds to the ratio between the effect-size of the association with resilience against AD (log of odds ratios of *C vs. P* comparison) and the effect-size of the association with AD (log of odds ratios of *A vs. P* comparison). We calculated the *change* in effect-size for the pPRS including and excluding *APOE* variants. We estimated 95% confidence intervals for the effect-size ratios by sampling, and we tested for significant difference between the *change* in effect-size including and excluding *APOE* variants (respectively for each of pPRS) using t-test. A value for the *change* in effect-size of 1 indicates a similar effect on increased risk of AD and resilience against AD. Although a value for the *change* in effect size unknown *a priori*, since all variants considered are selected to be associated with AD, a value <1 is expected (*i*.*e* a larger effect on AD than on resilience against AD).

### Contribution of each pathway to polygenic risk of AD

We estimated the contribution of each pathway to the genetic risk of AD in the general population: this equals to the variance explained by each of the pre-selected pathways to the genetic risk of AD. Mathematically, this is the ratio between the variance of each pathway-PRS and the variance of the combined PRS as calculated in the individuals in the general population. As such, it is a function of the *variant-pathway mapping*, the effect-size (log of odds ratio) of the variants, and the variant frequencies. Given a variant *k* and the relative *variant-pathway mapping* 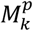, we define the percentage of the risk explained by each pathway *p* as:

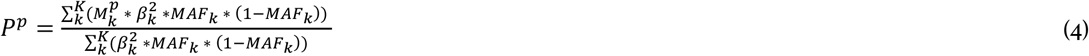

where *β*_*k*_ is the variant effect-size from literature, and *MAF*_*k*_ * (1 − *MAF*_*k*_) is the variance of a Bernoulli random variable that occurs with probability *MAF*_*k*_, *i*.*e* the minor allele frequency of each variant *k* in our cohort of population subjects. Here, 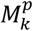 is interpreted as the probability that variant *k* belongs to pathway *p*. Importantly, for each variant, 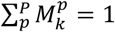, so that each variant contributes equally, yet differentially at the level of each pathway. This means that the variance of a variant is only counted once, even if the variant contributes to multiple pathways. When calculating the contributions of each pathway, we also considered variants with missing *variant-pathway mapping*. For these variants, the *variant-pathway mapping* was set to 1 for an *unmapped* pathway. Together, the pathway PRS variances sum to the total PRS variance.

### Implementation

We performed quality control of genotype data as well as population stratification analysis and relatedness analysis with PLINK (v2.0). All subsequent analyses were performed with R (v3.5.2), Bash and Python (v2.7.14) scripts. We provide a R script to construct pPRS and PRS using our *variant-pathway annotation* and user’s genotypes. In addition, all the scripts we used to perform the analyses can be found at https://github.com/TesiNicco/pathway-PRS.

## Results

After quality control of the genetic data, we included 1,654 population subjects (with mean age at inclusion 62.7±6.4, 53.2% females), 1,895 AD cases (with mean age at onset 69.2±9.9, 56.4% females), and 293 cognitively healthy centenarians (with mean age at inclusion 101.4±1.3, 72.6% females) (*P, A* and C respectively).

### Polygenic risk scores associate with AD and escape from AD

To each subject, we assigned a PRS representative of all 29 AD-associated variants, including and excluding *APOE* variants. We found that the PRS, when including *APOE* variants, significantly associated with an increased risk of AD and, in the opposite direction, with increased chance of resilience against AD (*A vs. P: OR=*2.61, 95% CI=[2.40-2.83], *p=*8.4×10^−113^ and *C vs. P: OR=*0.54, 95% CI=[0.45-0.65], *p=*1.1×10^−10^) (*Figure 1A* and *Table S11*). When excluding *APOE* variants, the PRS was still significantly associated with an increased risk of AD and, in the opposite direction, with increased risk of resilience against AD (*A vs. P: OR=*1.30, 95% CI=[1.22-1.40], *p=*3.1×10^−14^ and *C vs. P: OR=0*.*78*, 95% CI=[0.69-0.89], *p=*2.4×10^−4^) (*Figure 1B*, and *Table S11*).

**Figure 1:**
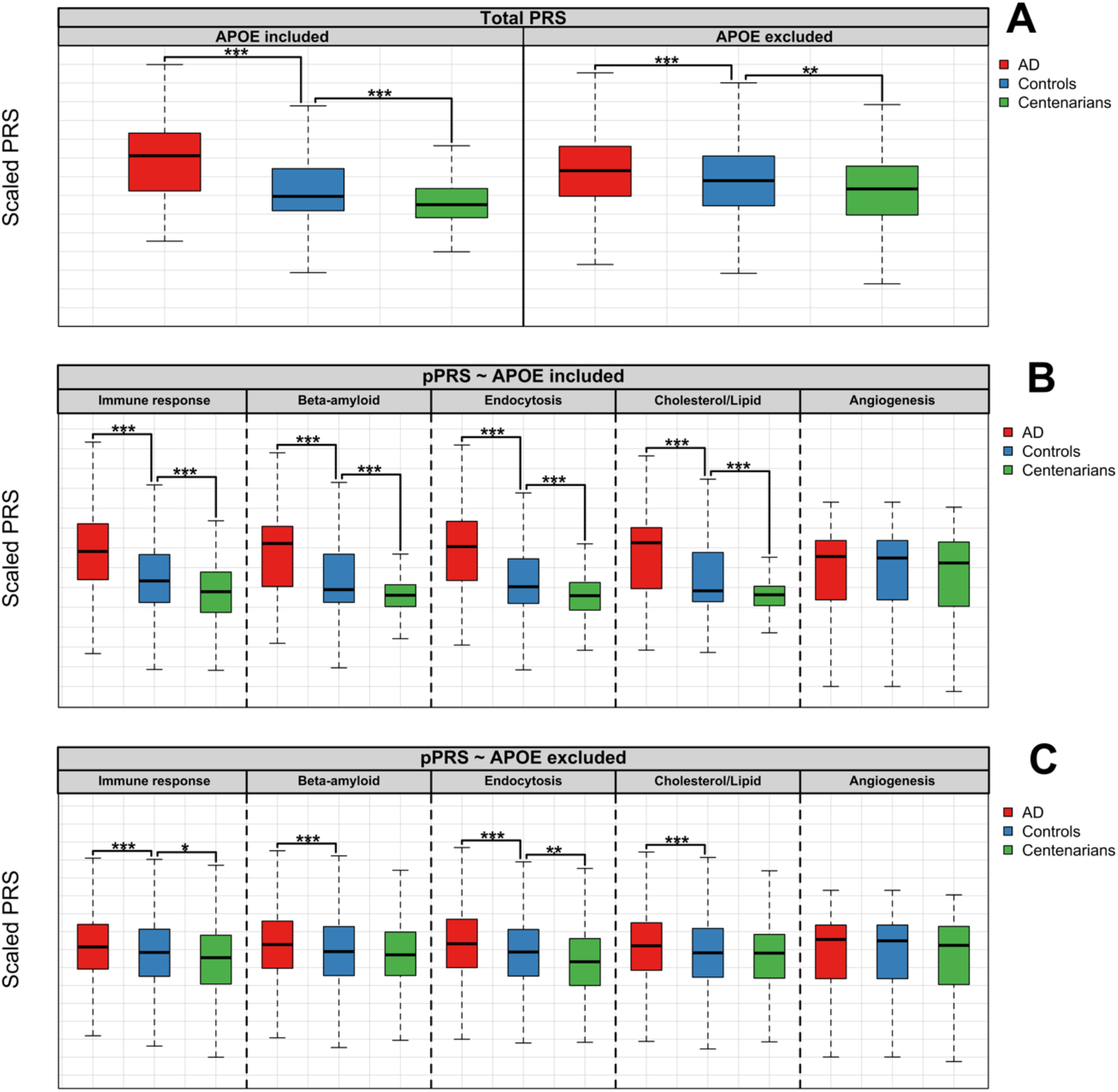
Boxplots of PRS and pPRS in the different settings. Figure A (above) shows the PRS including all the 29 known AD-associated variants, with and without *APOE* variants. As weight for the PRS, we used published variant effect-sizes (*Table S1*). Figure B (central) and Figure C (bottom) show the pPRS for each of the selected molecular pathways, including and excluding *APOE* variants, respectively. For all plots, risk scores were calculated for AD cases, population subjects and cognitively healthy centenarians. Then, risk scores were compared between (*i*) AD cases and population subjects (*A vs. P* comparison) and (*ii*) cognitively healthy centenarians and population subjects (*C vs. P* comparison). For representation, we scaled all PRS and pathway-PRS to be mean=0 and SD=1. For the comparison, we used logistic regression models with risk scores as predictors. Annotation: ***, *p-value* of association < 5×10^−6^; **, *p-value* of association < 5×10^−4^; *, *p-value* of association < 5×10^−2^.

### Pathway-specific PRS associate with AD and escape from AD

We annotated the 29 AD-associated genetic variants to 5 selected pathways *(Figure 2*). According to our *variant-gene mapping*, the 29 AD-associated variants mapped to 110 genes (*Table S8*). The number of genes associated with each variant ranged from 1 (*e*.*g*. for variants in/near CR1, *PILRA, SORL1, ABCA7, APOE* or *PLCG2*), to 30 (a variant in the gene-dense region within the *HLA* region) (*Figure 2* and *Table S8*). We were able to calculate the *gene-pathway mapping* weight for 69 genes (*Table S9*). The remaining 41 genes were not mapped to the 5 pathways. In total, we calculated the *variant-pathway mapping* for 23 loci to at least one of the pre-selected biological pathways (*Figure 2* and *Table S10*).

**Figure 2:**
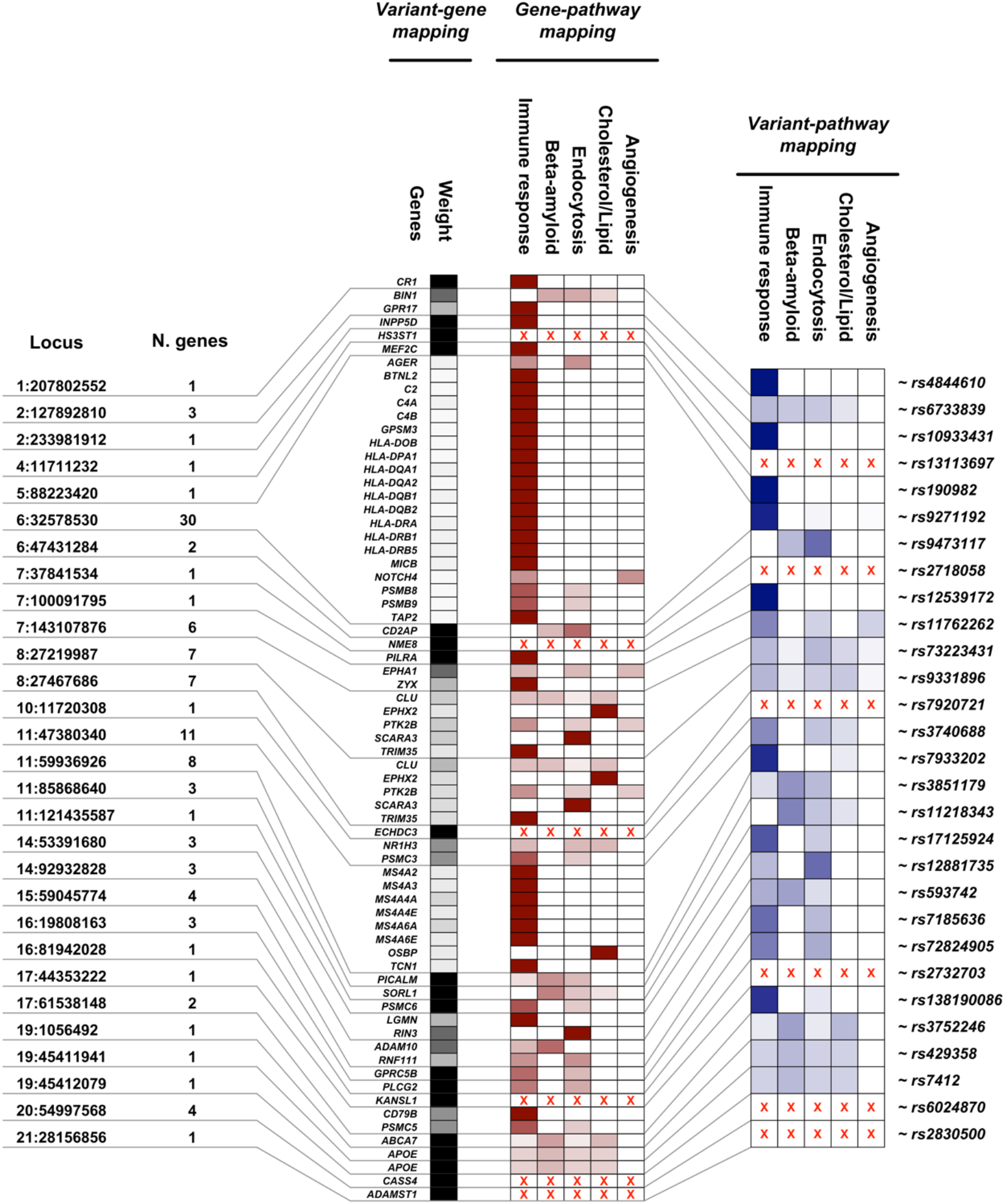
Variant-pathways mapping: *Locus*: chromosome and position of the AD-associated genetic variants (coordinates are with respect to GRCh37). *N. genes*: total number of genes associated with each variant according to *variant-gene mapping. Variant-gene mapping: Genes*: all genes with at least one annotation to the 5 selected molecular pathways associated with AD. *Weight*: the weight of the *variant-gene mapping. Gene-pathway mapping: Immune response, Beta-amyloid, Endocytosis, Cholesterol/lipid, Angiogenesis*: the weight of each molecular pathway at the gene level. *Variant-pathway mapping*: summarization of each variant’s effect after combining *variant-gene* and *gene-pathway* mappings. Red crosses indicate unmapped genes.

We then calculated the pPRS for each pathway in population subjects, AD cases and cognitively healthy centenarians including and excluding *APOE* variants (*Figure 1B* and *1C*). The number of variants that contributed to each pPRS was 19 for *immune response*, 11 for *β-amyloid metabolism*, 19 for *endocytosis*, 8 for *cholesterol/lipid dysfunction* and 4 for *angiogenesis* pathways (*Table S10* and *Table S11*). Overall, the pPRS (including and excluding the *APOE* variants) positively and significantly correlated with each other and with the overall PRS (*Figure S1*), and did not correlate with gender and age (*Figure S1*).

When including *APOE* variants, the pPRSs of all pathways (except for *angiogenesis*) significantly associated with increased risk of AD, independently from gender (*A vs. P, immune response*: *OR*=2.15, 95% CI=[1.99-2.32], *p*=2.0×10^−80^; *β-amyloid metabolism: OR=*2.52, 95% CI=[2.32-2.73], *p*=7.8×10^−109^; *endocytosis*: *OR*=2.55, 95% CI=[2.35-2.77], *p*=1.7×10^−109^; *cholesterol/lipid dysfunction*: *OR=*2.55, 95% CI=[2.35-2.76], *p*=2.1×10^−110^; *angiogenesis: OR*=1.05, 95% CI=[0.98-1.12], *p*=0.134) (*Figure 1B, Table S11, Figure S2* and *Table S12*). The association of pPRSs with increased chance of being resilient against AD was in the opposite direction for all pathways, and the association was significant for all pathways except for *angiogenesis* (*C vs. P, immune response*: *OR*=0.64, 95% CI=[0.54-0.74], *p*=1.4×10^−8^; *β-amyloid metabolism: OR*=0.59, 95% CI=[0.49-0.71], *p*=2.7×10^−8^; *endocytosis*: *OR*=0.55, 95% CI=[0.46-0.66], *p*=1.3×10^−10^; *cholesterol/lipid dysfunction: OR*=0.58, 95% CI=[0.48-0.70], *p*=1.8×10^−8^; *angiogenesis: OR*=0.90, 95% CI=[0.79-1.01], *p*=0.078) (*Figure 1B, Table S11*). Directions of effects were consistent in both males and females, but the significance of associations was reduced due to stratification (*Table S12* and *Figure S2*).

When excluding *APOE* variants, the pPRSs of all pathways (except for the *angiogenesis*) was still significantly associated with increased risk of AD without specific gender effects (*A vs. P, immune response*: *OR*=1.19, 95% CI=[1.11-1.27], *p*=5.5×10^−7^; *β-amyloid metabolism: OR=*1.19, 95% CI=[1.12-1.28], *p*=2.0×10^−7^; *endocytosis*: *OR*=1.27, 95% CI=[1.19-1.36], *p*=2.8×10^−12^; *cholesterol/lipid dysfunction*: *OR=*1.18, 95% CI=[1.11-1.27], *p*=7.5×10^−7^; *angiogenesis: OR*=1.05, 95% CI=[0.98-1.12], *p*=0.134) (*Figure 1C, Table S11, Figure S2* and *Table S12*). The association of pPRSs with increased chance of being resilient against AD was in the opposite direction for all pathways, yet the association was significant only for the *immune response* and the *endocytosis* pPRS (*C vs. P, immune response*: *OR*=0.82, 95% CI=[0.72-0.94], *p*=0.003; *β-amyloid metabolism: OR*=0.91, 95% CI=[0.80-1.03], *p*=0.131; *endocytosis*: *OR*=0.79, 95% CI=[0.70-0.90], *p*=2.8×10^−4^; *cholesterol/lipid dysfunction: OR*=0.91, 95% CI=[0.80-1.03], *p*=0.145; *angiogenesis: OR*=0.90, 95% CI=[0.79-1.01], *p*=0.078) (*Figure 1C* and *Table S11*). In the sex-stratified analysis, females reported consistent direction of effects and significant associations of *immune response* and *endocytosis* pathways, while in males the direction was consistent for *immune response, endocytosis* and *angiogenesis* pathways, and it was opposite for *β-amyloid metabolism* and *cholesterol/lipid dysfunction* (yet not significant) (*Figure S2* and *Table S12*).

We note that apart from *APOE* variants (for which we stratified the analyses for), there was no major driver in the pPRS as well as the single-variant associations (*Figure S3* and *Figure S4*).

### Comparison of effect on AD and escaping AD

To further evaluate the association of the pPRSs with AD and with resilience against AD, we compared, for each pPRS, the reciprocal effect size associated with resilience against AD with the effect size associated with increased risk of AD (*change* in effect size, *Figure 3A*). When including *APOE* variants, the *change* in effect-size was <1 for all pathways (except for the *angiogenesis* pathway) (*Figure 3B*). This is expected as the effect-size of *APOE* variants on *causing* AD is much larger than its effect on resilience against AD (*Figure 3A*). When excluding *APOE* variants, the *change* in effect-size was still <1 for *β-amyloid metabolism* and *cholesterol/lipid metabolism* (respectively 0.54 and 0.58), but it approximated 1 for *endocytosis* (0.96) and it was larger than 1 for the *immune response* and *angiogenesis* (respectively 1.12 and 2.15) (*Figure 3B*). Interestingly, we found that the relative effect-size for *immune response* and *endocytosis* excluding *APOE* variants was significantly higher than that including *APOE* variants (*p*<2.1×10^−197^ and *p*<8.9×10^−180^ respectively), suggesting a larger effect on resilience against AD compared to AD-risk for these pathways, specifically when excluding *APOE* variants (*Figure 3B*).

**Figure 3:**
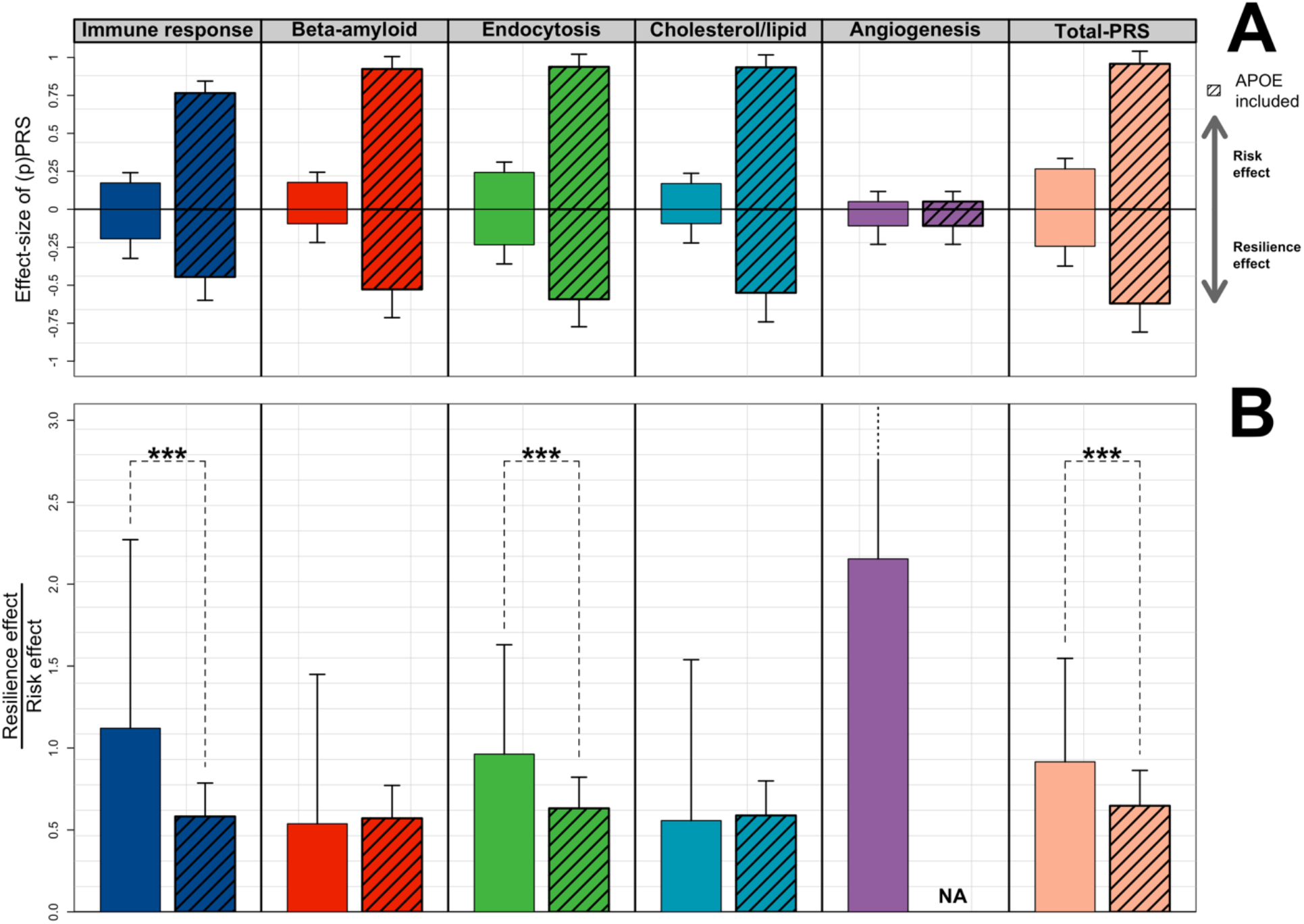
*Change* in effect-size between association with escaping AD and causing AD for the five pPRSs: figure A shows the effect-sizes (log of odds ratio) and the relative 95% confidence intervals of the association of the (p)PRS with both AD-risk and resilience against AD, grouped by pathway. In figure B, each bar represents the ratio between the effect-size of the association with escaping AD (*Resilience effect* in figure A) and with causing AD (*Risk effect* in figure A), respectively with and without *APOE* variants. Ratios larger than 1 are then indicative of larger effect-size on resilience against AD compared to AD-risk. We then compared the *change* in effect-size for each pathway when including and excluding *APOE* variants using t-tests. Annotation: ***, *p-value* of association < 5×10^−6^; **, *p-value* of association < 5×10^−4^; *, *p-value* of association < 5×10^−2^.

### Contributions of each pathway to the polygenic risk of AD

Finally, we estimated the relative contribution of each pathway to the polygenic risk of AD in the general population. This is indicative of the degree of involvement of each pathway to the total polygenic risk of AD, and as such it is based on out *variant-pathway mapping*. Including *APOE* variants, the contribution of the pathways to the total polygenic risk of AD was 29.6% for *β-amyloid metabolism, 26*.*6%* for *immune response*, 21.6% for *endocytosis*, 19.5% for *cholesterol/lipid dysfunction, 0*.*3%* for *angiogenesis* and 2.3% for the *unmapped* variants (*Figure 4A*).

**Figure 4:**
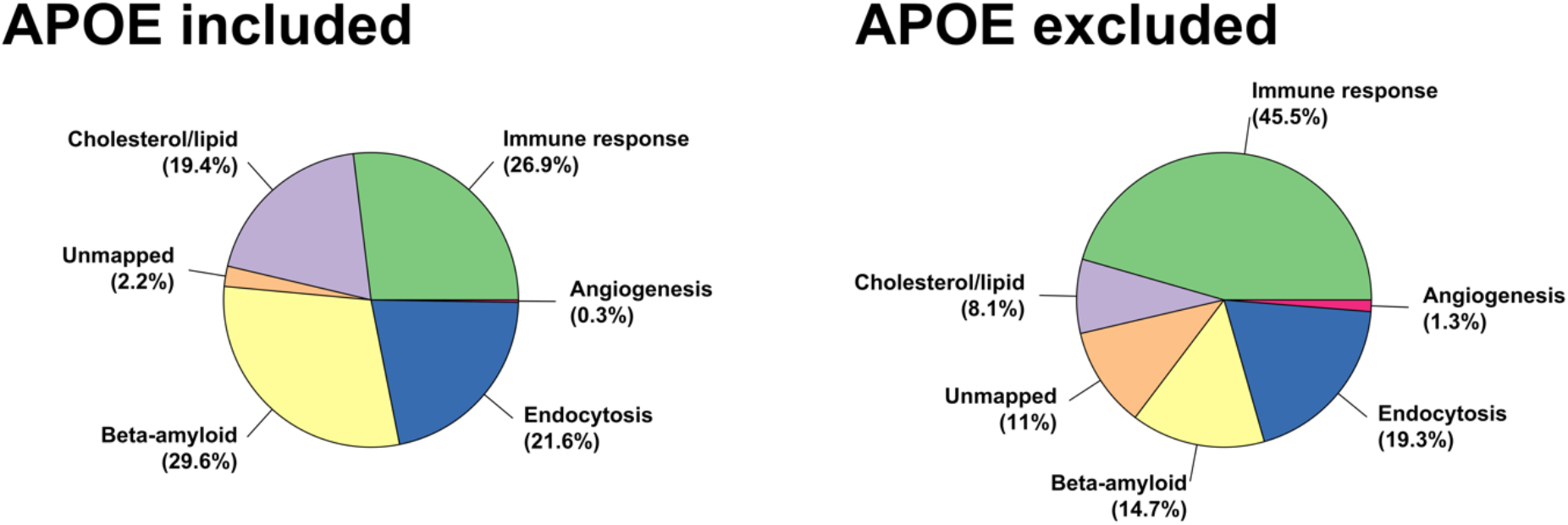
Explained variance of each pathway-specific PRS to polygenic risk of AD: the pie charts represents the explained variance of each pathway-specific PRS to the polygenic risk of AD, including and excluding *APOE* variants. The contributions are calculated according to (*i*) our *variant-pathway mapping*, (*ii*) the effect size (log of odds ratio) of each variant from literature (*Table S1*), and (*iii*) variant’s frequency in our cohort of middle-aged healthy population subjects. We also considered variants with missing *variant-pathway mapping* (*unmapped* pathway).

When we excluded *APOE* variants, the contribution of the pathways to the total polygenic risk of AD was 45.5% for *immune response*, 19.2% for *endocytosis*, 13.7% for *β-amyloid metabolism*, 8% for *cholesterol/lipid dysfunction*, 1.4*%* for *angiogenesis* and 12.3% for the *unmapped* variants (*Figure 4B*).

## Discussion

In this work, we studied 29 common genetic variants known to associate with AD using polygenic risk scores and pathway-specific polygenic risk scores. As expected, we found that a higher PRS for AD was associated with a higher risk of AD. Previous studies showed that polygenic risk score of AD not only associated with increased risk of AD, but also with neuropathological hallmarks of AD, lifetime risk and the age at onset in both *APOE ε4* carriers and non-carriers.[28, 29, 51–55] We now add that, using our unique cohort of cognitively healthy centenarians, the PRS for AD also associates with resilience against AD at extremely old ages. This adds further importance to the potentiality of using PRS and *APOE* genotype in a clinical setting.[51, 52, 54, 56] In addition, our analyses suggest that the long-term preservation of cognitive health is associated with the selective survival of individuals with the lowest burden of risk-increasing variants or, vice versa, the highest burden of protective variants.

Using an innovative approach, we studied five pathways previously found to be involved in AD as well as the contribution of these pathways to the polygenic risk of AD. We showed that all pathways-PRS except *angiogenesis* associate with increased AD risk, both including and excluding *APOE* variants and independently from gender. When we studied the association of pathways-PRS with resilience against AD until extreme old ages, we found that, as expected, the enrichment of the protective *APOE ε2* allele and the depletion of the risk-increasing *APOE ε4* allele represented a major factor in avoiding AD. However, when excluding the two *APOE* variants, only *immune response* and *endocytosis* significantly associated with an increased chance to be resilient against AD. Interestingly, both pathways had a larger or similar effect on resilience against AD-resilience compared to developing AD, suggesting that these pathways might be involved in general neuro-protective functions. Based on the variant effect size, variant frequency and our *variant-pathway mapping*, we found that the *β-amyloid metabolism* (29.6%) followed by *immune response* (26.6%) were the major contributors to general modification of AD-risk. After excluding *APOE* variants, according to our analysis, *immune response* (45.5%) and *endocytosis* (19.2%) contributed most to the modification of AD-risk.

Our approach to map variants to associated genes and to map genes to pathways resulted in a weighted annotation of variants to pathways that allowed for uncertainty in gene as well as pathway assignment, which was not done previously. We note that considering uncertainty in variant-gene as well as gene-pathway assignments is crucial because most genetic variants are in non-coding regions, which makes the closest gene not necessarily the culprit gene, and because different functional annotation-sources often do not overlap. In our *variant-pathway mapping*, a larger number of annotations (both variant-genes and gene-pathways), generally causes a dilution of the “true” variant effect, reflecting increasing uncertainty in the annotation sources used. This depends on the specific regions, for example, the HLA region carries many genes with large linkage signals, however, all genes in this region are typically annotated with immune response. We point out that the power of the PRSs does not only reflect the effect-size of the variants, but also the number and frequency of the variants that contribute to the PRSs: due to this, a larger number of very common variants with relatively small effect-size can still have more power (yet small ORs) than a small number of relatively rare variants with high effect-size. The pathway-specific PRS that we proposed in this manuscript can be re-used for the identification of subtypes of AD patients compromised in a specific AD-associated pathway. This is of interest for clinical trials, in order to test responsiveness to compounds in specific subsets of patients. For example, monoclonal antibody targeting *TREM2* receptors could work better in AD patients who have an impaired immune response pathway. Recently, several studies attempted to construct pathway-specific PRS to find heterogeneity in AD patients based on a genetic basis.[28, 29] In line with our findings, *Ahmad et al*. found that genes capturing *endocytosis* pathway significantly associated with AD and with the conversion to AD.[29] Other studies used less variants [28] or less stringent selection for variants, and did not observe a differential involvement of pathways in AD etiology.[57]

The amyloid cascade hypothesis has been dominating AD-related research in the last two decades. However, treatments targeting amyloid have, so far, not been able to slow or stop disease progression. This has led to an increased interest for the other pathways that are important in AD pathogenesis.[22] Part of the current view of the etiology of AD is that the dysregulation of the immune response is a major causal pathway, and that AD is not only a consequence of β-amyloid metabolism.[58, 59] In addition, previous studies showed that healthy immune and metabolic systems are associated with longer and healthier lifespan.[1, 60] Our results indicate that, excluding *APOE* variants, the effect of immune response and endocytosis on escaping AD is stronger or comparable to the effect on causing AD. This suggests that these pathways might be involved in the maintenance of general cognitive health, as the cognitively healthy centenarians represent the escape of all neurodegenerative diseases until extreme ages. We recently found evidence for this hypothesis in the protective low frequency variant in *PLCG2*, which is involved in the regulation of the immune response.[53] This variant is enriched in cognitively healthy centenarians, and protects against AD as well as frontotemporal dementia and dementia with Lewy bodies.[53] We included this variant in the total PRS as well as in the pathway-PRS for the immune response (*variant-pathway mapping* was 60%) and endocytosis (*variant-pathway mapping* was 40%). Regarding endocytosis, this pathway is thought to play a role both in neurons, as part of the β-amyloid metabolism, but also in glia cells, as part of the immune response. Thus, a dysregulation in the interplay between these pathways might lead to an imbalance of immune signaling factors, favoring the engulfment of synapses and AD-associated processes. This, in turn, may contribute to the accumulation of amyloid and tau pathologies.[61–64].

We assessed the effect of common and low frequency variants on the development and the escape of AD. Therefore, the contributions of rare, causative variants associated with increased AD risk, such as those in *APP, PSEN1, PSEN2, TREM2* and *SORL1* were not considered. Despite the large odds ratios to develop AD associated with carrying such variants, the frequency of these variants in the population is ultra-low, and therefore have a minor effect on the total AD risk in the population.[11, 12] However, future versions of the PRS will most likely include the effect of carrying disease-associated rare variants. This will affect individual PRS scores and the necessity to accordingly adapt the results generated with current PRSs. Compared to the sizes of recent GWAS of AD, we included relatively small sample sizes, particularly with respect to the cognitively healthy centenarians, a very rare phenotype in the population (<0.1%).[4] These sample sizes are however sufficient to study PRSs. The cohorts that we used in this study were not used in any GWAS of AD, therefore we provide independent replication of AD PRS in a homogeneous group of (Dutch) individuals.

We note that, apart from *APOE* variants (for which we stratify the analyses for), none of the other variants have been associated with longevity or well cognitive functioning in the largest and most recent GWAS.[65, 66] We acknowledge that our *variant-pathway mapping* reflects the current state of imperfect knowledge at the level of AD-GWAS findings, variant-gene and gene-mechanism relationships. Thus, as new variants, pathways or functional relationships will be identified, the contributions and the pathway-specific PRSs will need to be recalculated. Of note: the study in which the genome-wide significant association with AD of the variant in/near *KANSL1* was originally identified, reported a larger effect size compared to the effect size used in our manuscript, (β=0.31 and β=0.07, respectively), possibly because the original analysis was stratified by *APOE*. We cannot exclude that we underestimated the contribution of *KANSL1* in the analyses. Moreover, since the *KANSL1* variant did not map into one of the analyzed pathways, it was not included in any of the pathway-specific PRS calculations. A limitation, not exclusive to our work, is the highly debated role of *APOE* gene. We mapped the effect of *APOE* to four pathways and we are aware this assignment is relatively arbitrary. We add that *APOE* has well-studied (cardio)vascular properties that are included in our cholesterol and lipid metabolism pathway. The combination of a large effect and unclear pathway assignment makes that pathway-PRS including *APOE* challenging to use. Lastly, we point out that the variance contributions might change in different populations, as it depends on variant frequency and population heterogeneity.

Concluding, with the exclusion of *APOE* variants and based on our functional annotation of variants, the aggregate contribution of the immune response and endocytosis represents more than 60% of the currently known polygenic risk of AD. This indicates that an intervention in these systems may have large potential to prevent AD and potentially other related diseases and highlights the critical need to study (neuro)immune response and endocytosis, next to β-amyloid metabolism.

## Data Availability

We provide in the supplements the estimates to construct pathway-specific polygenic risk scores with any genotype data. The genotype data that we used in this work are not publicly available. If interested, you may contact us.

https://github.com/TesiNicco/pathway-PRS

## Acknowledgements

The following studies and consortia have contributed to this manuscript. Amsterdam dementia cohort (ADC): Research of the Alzheimer center Amsterdam is part of the neurodegeneration research program of Amsterdam Neuroscience (www.amsterdamresearch.org). The Alzheimer Center Amsterdam is supported by Stichting Alzheimer Nederland and Stiching VUmc fonds. The clinical database structure was developed with funding from Stichting Dioraphte. Genotyping of the Dutch case-control samples was performed in the context of EADB (European Alzheimer DNA biobank) funded by the JPco-fuND FP-829-029 (ZonMW projectnumber 733051061). 100-plus study: we are grateful for the collaborative efforts of all participating centenarians and their family members and/or relatives. This work was supported by Stichting Alzheimer Nederland (WE09.2014-03), Stichting Diorapthe, horstingstuit foundation, Memorabel (ZonMW projectnumber 733050814) and Stichting VUmc Fonds. Genotyping of the 100-plus study was performed in the context of EADB (European Alzheimer DNA biobank) funded by the JPco-fuND FP-829-029 (ZonMW projectnumber 733051061). The clinical database structure was developed with funding from Stichting Dioraphte. Longitudinal Aging Study Amsterdam (LASA) is largely supported by a grant from the Netherlands Ministry of Health, Welfare and Sports, Directorate of Long-Term Care. The authors are grateful to all LASA participants, the fieldwork team and all researchers for their ongoing commitment to the study. This work was in part carried out on the Dutch national e-infrastructure with the support of SURF Cooperative. The authors declare no conflict of interests.

